# COVID-19 Projections for K12 Schools in Fall 2021: Significant Transmission without Interventions

**DOI:** 10.1101/2021.08.10.21261726

**Authors:** Yiwei Zhang, Karl Johnson, Zhuoting Yu, Akane B. Fujimoto, Kristen Hassmiller Lich, Julie Ivy, Pinar Keskinocak, Maria Mayorga, Julie L. Swann

**Affiliations:** Department of Industrial and Systems Engineering, North Carolina State University, Raleigh, NC, United States of America; Department of Health Policy and Management, University of North Carolina at Chapel Hill, Chapel Hill, NC, United States of America; Department of Industrial and Systems Engineering, Georgia Institute of Technology, Atlanta, GA, United States of America

## Abstract

**Background:** Millions of primary school students across the United States are about to return to in-person learning. Amidst circulation of the highly infectious Delta variant, there is danger that without the appropriate safety precautions, substantial amount of school-based spread of COVID-19 may occur.

**Methods:** We used an extended Susceptible-Infected-Recovered computational model to estimate the number of new infections during 1 semester among a student population under different assumptions about mask usage, routine testing, and levels of incoming protection. Our analysis considers three levels of incoming protection (30%, 40%, or 50%; denoted as “low”, “mid”, or “high”). Universal mask usage decreases infectivity by 50%, and weekly testing may occur among 50% of the student population; positive tests prompt quarantine until recovery, with compliance contingent on symptom status.

**Results:** Without masking and testing, more than 75% of susceptible students become get infected within three months in all settings. With masking, this values decreases to 50% for “low” incoming protection settings (“mid”=35%, “high”=24%). Testing half the masked population (“testing”) further drops infections to 22% (16%, 13%).

**Conclusion:** Without interventions in place, the vast majority of susceptible students will become infected through the semester. Universal masking can reduce student infections by 26-78%, and biweekly testing along with masking reduces infections by another 50%. To prevent new infections in the community, limit school absences, and maintain in-person learning, interventions such as masking and testing must be implemented widely, especially among elementary school settings in which children are not yet eligible for the vaccine.

## Introduction

During summer 2021, the Delta variant of SARS-CoV-2 has spread with a substantially higher infectivity than previous strains. Millions of students across the US are scheduled to soon return to K12 classrooms, and vaccination among them is limited (middle and high school) or unavailable (elementary school). Policymakers, school systems, and parents are evaluating the potential impact of school openings and interventions such as masks and regular testing in K12 schools on the spread of SARS-CoV-2. This analysis assesses the benefits of layered interventions in schools.

## Methods

An extended Susceptible-Infected-Recovered model was developed using R programming language to project infections over one semester (107 days) within a well-mixed student population (n=500) in which 0.5% of incoming students are infected and one case enters the school per week (e.g., infected outside school). Weekly RT PCR testing of 50% of students with sensitivity (specificity) of 85% (95%) is assessed. Students who don’t receive testing are never put in isolation, regardless of disease status. Students who receive testing but are uninfected are also never put in isolation. However, those who are infected and receive testing have a high probability of being put in isolation with compliance (70%, 90%) contingent on symptoms (asymptomatic, symptomatic); these students remain in testing until they are fully recovered. Infected students who test positive and are isolated can no longer infect susceptible students. Alternatively, infected students who do not test positive have the capacity to continuously infect other students until they eventually recover. Testing is random among members of the student population that have not already recovered and are not in isolation. Case-investigation of potential contacts is not conducted. We modeled three different school settings across different levels of incoming protection (through natural or vaccine-acquired immunity) of 30%, 40%, or 50%; for convenience these settings are denoted as “low”, “mid”, or “high”, reflecting different communities or grade levels (i.e., “low” = elementary school, ‘mid’ or ‘high’ reflecting middle or high school). Levels of protection were based on CDC reports that 30% of students in the middle school age-range are vaccinated, 40% of students in the high-school age-range are vaccinated, and prior infection among all primary-school children is approximately 10%.^1^ A baseline effective reproductive rate (R_0_) of 4.0 represented the context of increased infectivity from the Delta variant. We assume universal mask usage decreases infectivity by 50%.^2^ A scenario analysis considered an R_0_ of 5.0. All model parameters are provided in Table 1.

**Table 1.** Model Parameters and Values.

| Table 1 - Model Parameters and Values |  |  |
| --- | --- | --- |
| Parameters | Description | Value |
| T | Number of days modeled | 107 |
| ppl_size | School size | 500 |
| prob0 | Proportion of infections that are symptomatic | 0.6 |
| prevalence | Initial infections within the population | 0.005 |
| i_new | New infections per week | 1 |
| recovered | Incoming protection | 0.3, 0.4, 0.5 |
| test_level_pcr | Testing level of PCR test | 0, 0.5 |
| test_level_rap | Testing level of Rapid test | 0 |
| $\beta_a$ , $\beta_s$ | Transmission rate of due to contact between susceptible and asymptomatic/symptomatic infected individuals | (0.125, 0.25), (0.25, 0.5) |
| $\gamma_a$ , $\gamma_s$ | Rate of recovery of an asymptomatic/symptomatic infected individuals | 0.1, 0.1 |
| $\mu$ | Mortality rate | 0 |
| Ca, Cs | Isolation compliance rate for asymptomatic/symptomatic infected and tested individuals | 0.7, 0.9 |
| FNpcr,a , FNpcr,s | False negative rate of PCR test when individual is asymptomatic/symptomatic | 15%, 15% |
| FNrap,a , FNrap,s | False negative rate of rapid test when individual is asymptomatic/symptomatic | 30%, 30% |
| FPpcr,a , FPpcr,s | False positive rate of PCR test when individual is asymptomatic/symptomatic | 5%, 5% |
| FPrap,a , FPrap,s | False positive rate of PCR test when individual is asymptomatic/symptomatic | 10%, 10% |

## Results

Without testing and masking (R_0_=4.0), more than 75% of susceptible students get infected within three months in all settings (Figure 1). With masks (R_0_=2.0), the proportion infected drops to approximately 50% for “low” incoming protection settings (“mid”=35%, “high”=24%). Testing half the masked population (“testing”) further drops infections to 22% (16%, 13%). If R_0_ is 5.0, 95% (93%, 88%) can be infected without mitigation and 70% (57%, 41%) with masking for the “low” (“mid”, “high”) setting. Assuming a conservative total of 10 days of school absence per 5 new infections, there will be an estimated 210 (510, 400) absent days for the school without any interventions or 140 (120, 76) days with masking and testing.

**Figure 1:**
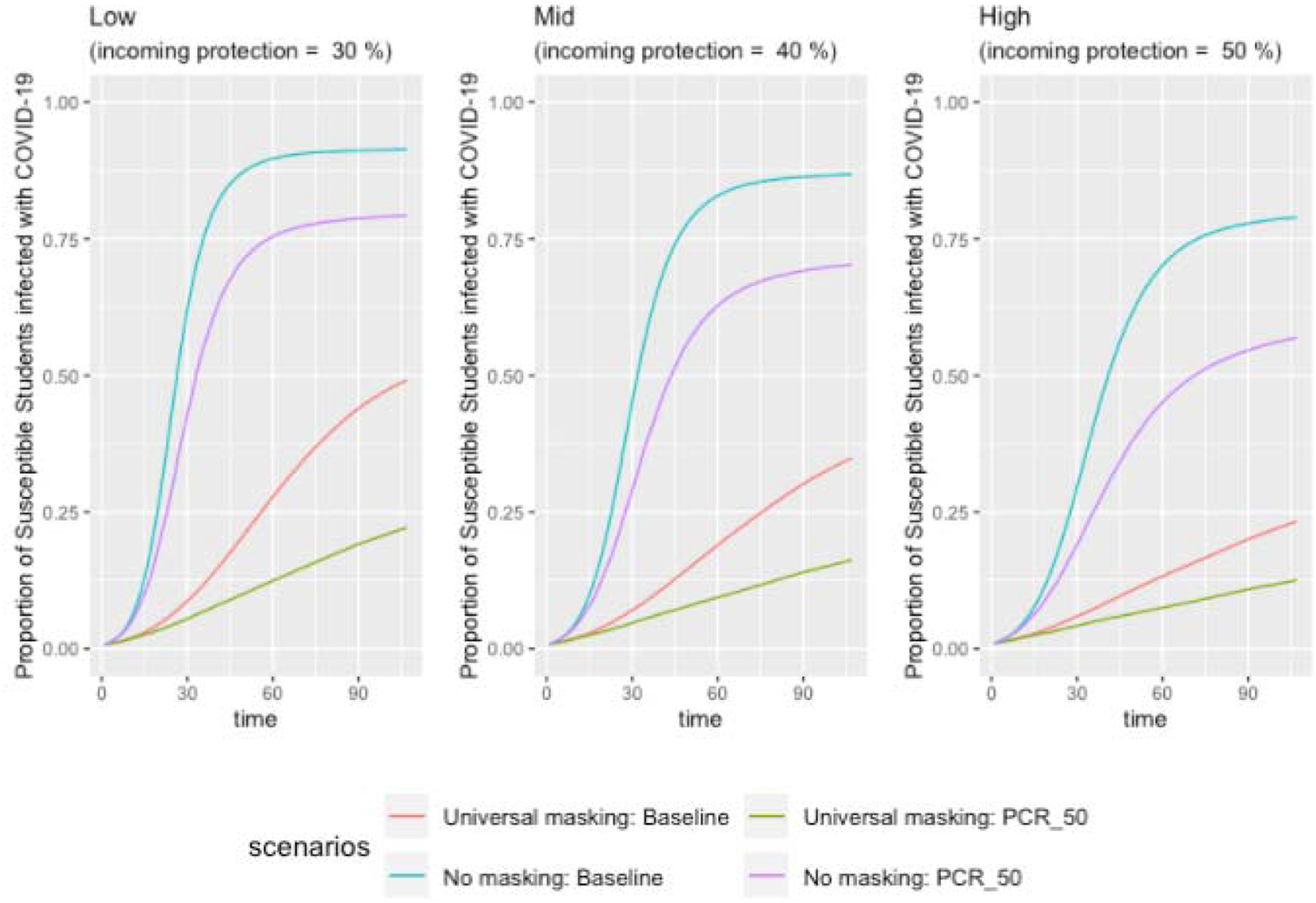
Projected Spread of COVID-19 across Epidemiological Setting and School Policies: Each of the following panels presents the spread within the three different epidemiological settings of our model (“Low” = incoming protection of 30%, “Mid” = incoming protection of 40%, “High” = incoming protection of 50%). The Y axis represents the fraction of susceptible students who become infected throughout the semester (107 days). The “No Masking” scenarios reflect disease spread with an effective reproductive rate of four and the “Universal Masking” scenarios reflect an effective reproductive rate of 2, representing a 50% reduction in transmission from masking. Across each setting and masking scenario, two different testing policies are modeled: either no testing (“baseline”) or randomly testing half of students every week (“PCR_50”).

**Figure 2.**
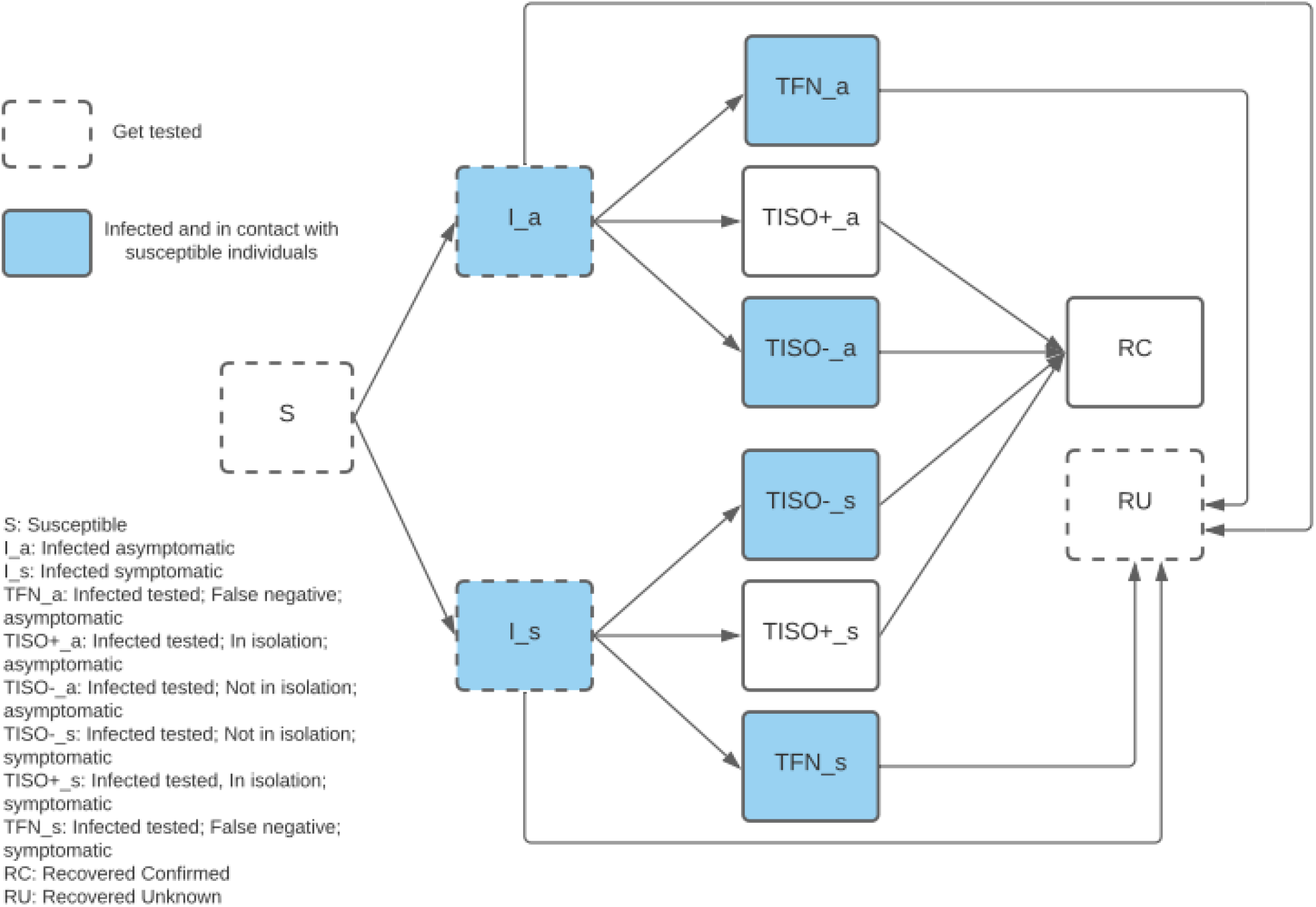
Testing and Isolation Pathways within Extended SIR Model: Within our model, various pathways are followed among students who received testing.

## Discussion

Without interventions in place, the vast majority of susceptible students among K12 schools will become infected and school absences will increase, followed by additional cases in communities as infected students transmit to household members. Universal masking can reduce student infections by 26-78%, and testing biweekly along with masking reduces infections by another 50%. Self-quarantine among exposed students or virtual-by-choice options may further reduce infections.^3^ Interventions should remain until most students are fully vaccinated. Jurisdictions need time to reduce inequities in vaccine uptake, and they may periodically need interventions when community spread is high.

The link between Delta and long-Covid or multi-inflammatory syndrome is unknown, and the risk of the most severe disease is lower among children but may exist.^4,5^ However, there are mental health concerns,^6^ learning gaps,^7^ and other implications if schools remain closed. The multitude of direct public health and indirect social and economic benefits make layered interventions worth the investment.

## Data Availability

All data utilized for the development of this model is available upon request.

